# “Maternal and Socioeconomic Factors Drive Childhood Anemia in Tajikistan: Examining the Role of Zero Vegetable or Fruit Consumption among under five”

**DOI:** 10.1101/2025.10.24.25338624

**Authors:** Asif Khaliq, Namra Ijaz, Yusra Rizwan, Sarah Aijaz, Bushra Ashar

## Abstract

**Objectives:** To examine whether complete absence of vegetable and fruit consumption (Zero Vegetable or Fruit - ZVF) independently associates with anemia among children aged 0-59 months in Tajikistan, and to identify key maternal and socioeconomic determinants of childhood anemia.

**Design:** Cross-sectional analysis using recent 2023, nationally representative Demographic and Health survey data from Tajikistan.

**Setting:** National population based study across all regions of Tajikistan (urban and rural areas) using two stage stratified cluster sampling.

**Participants:** 2,355 children in age group 0-59 months with complete data on hemoglobin levels and dietary intake in the preceding 24 hours residing in households selected from 2023 Tajikistan Demographic and Health Survey were included in this study.

**Main Outcome Measures:** Primary outcome, childhood anemia status (hemoglobin <10.5g/dl for 6-23 months, <11g/dl for 24-59 months, altitude-adjusted per WHO 2024 criteria).

**Results:** Among Tajik children under 5 years, 36.1% of children consumed no vegetables or fruits in the preceding 24 hours and the prevalence of anemia was 33.2%. Adjusted odds in our study revealed no association of ZVF consumption with anemia (OR=0.85, 95% CI: 0.63 to 1.14). Significant associations emerged for maternal anemia which increased child anemia odds by 48% (OR=1.48, 95% CI: 1.16 to 1.88), richest index had 60% lower anemia odds compared to the poorest (OR=0.39, 95% CI: 0.27 to 0.56) and low birth size increased anemia risk two fold (OR=1.92, 95% CI: 1.31 to 2.81).Each additional daily meal consumption reduced anemia odds by 14% (OR=0.86, 95% CI: 0.76 to 0.98).

**Conclusions:** Child anemia in Tajikistan is not primarily driven by specific dietary factors but maternal nutritional status during pregnancy, birth outcomes, and socioeconomic disparities are main drivers. This novel null finding on ZVF intake, underscores that non dietary drivers predominate, informing targeted interventions aligned with SDG2 (Zero hunger). Thus integrated approaches aim to address maternal nutrition, antenatal care, wealth inequalities, and feeding frequency promotion.

**What is already known on this topic:** Over 40% of children under five globally are affected by childhood anemia with dietary factors like low fruit and vegetable intake considered important contributors. However, the specific impact of complete absence (zero consumption) of vegetables and fruits on childhood anemia needs to be explored particularly in Central Asian countries like Tajikistan as here the dietary pattern differ from other low- and middle-income settings.

**What this study adds:** This first nationally representative Central Asian study showed that downstream factor that’s the zero vegetable or fruit consumption had no association with childhood anemia in Tajikistan. Instead upstream factors like maternal anemia during pregnancy, household wealth disparities, small birth size, and meal frequency emerged as primary determinants, challenging dietary-centric intervention approaches.

**How this study might affect research, practice or policy:** These findings redirect anemia prevention strategies towards integrated interventions addressing maternal nutrition during pregnancy, poverty alleviation, improved antenatal care for fetal growth monitoring, and feeding frequency promotion rather than focusing solely on vegetable or fruit consumption. This evidence informs sustainable development goals (SDG) aligned nutrition policies requiring upstream determinants to be prioritized alongside dietary diversity programs in similar settings.

**Key Points:** *Core Issues:* This study explored whether complete absence of vegetable and fruit consumption (ZVF) associates with childhood anemia (below five years) in Tajikistan thereby challenging dietary-centric intervention approaches by examining maternal and socioeconomic determinants.

*Findings:* The research found no significant association between zero vegetable or fruit consumption with childhood anemia, but identified maternal anemia during pregnancy, household wealth disparities, small birth size, and meal frequency as primary drivers.

*Meaning:* This novel null finding of our research redirects anemia prevention strategies from dietary diversity alone toward integrated upstream interventions addressing maternal nutrition, poverty alleviation, antenatal care quality, and feeding frequency informing SDG-aligned policies in Central Asia and similar Lower-and Middle-income countries.

## Introduction

Anemia remains a major public health concern especially for children of age 6-59 months. According to the estimates of World Health Organization (WHO) 2025 showed over 40% of children under five years of age worldwide to be affected(1). Especially a concern in rural areas some related factors like limited dietary diversity, food insecurity, seasonal poverty, and inadequate access to nutritious food such as iron-fortified products, fruits, vegetables, and animal proteins,there by prevalence is consistently rising(2). The weaning period is the time of maximum burden which a child under 2 years face in low- and middle-income countries as evidenced by the United Nations Children’s Fund (UNICEF)(3) and also reports the intergenerational impacts of child anemia with maternal anemia(4). During infancy and early childhood, stunted growth, decreased immunity, and poor cognitive and psychomotor development are heavily influenced by anemia(5). These effects not only are deleterious for child survival but also compromise long-term educational achievement and productivity, leading to vicious cycles of poverty and ill health. Anemia caused ∼52 million years lived with disability (YLDs) globally in 2021, with under 5s contributing 20%(6). While SDG 2.2 anemia target focuses on women, interventions for children under 5 are critical to achieving broader malnutrition goals, as maternal and child anemia are interlinked(3). The stable 40% prevalence (∼269 million children in 2025)from WHO/UNICEF estimates underscores the urgency of scaling up interventions (4)to meet the SDG target 2030.

Fruits and vegetables are rich sources of non-heme iron, vitamin C, folate, and antioxidants, all crucial for hemoglobin synthesis and red blood cell production. Although non-heme iron from plant sources is less bioavailable than heme iron from animal foods, vitamin C in fruits and vegetables enhances its absorption by reducing ferric (Fe^3+^) to ferrous (Fe^2+^) iron(7). The Zero Vegetable or Fruit consumption (ZVF) a new standard variable was introduced in the 2021 update to the Infant and Young Child Feeding (IYCF) guidelines by the WHO & UNICEF, 2021 (8)for household surveys to measure unhealthy eating practices. The target children for this variable are aged 6–23 months who did not consume vegetables or fruits in the previous day or night. This indicator is critical because inadequate vegetable and fruit intake heighten anemia and some studies support stunting risk (9) and in long-term leads to heart disease and type 2 diabetes (8).

Tajikistan representing a LMIC in Central Asia, shows similar burden moreover where rural households often rely on self grown staples like cereals and potatoes, which are low in bioavailable iron, exacerbating micronutrient deficiencies(10). While global evidence relates low fruit and vegetable consumption to anemia, research on the impact of entire absence (“zero consumption”) remains limited, and no earlier study has specifically addressed this association in Tajikistan(11,12). This represents a critical regional gap, as Central Asia specific studies on dietary anemia specifically ZVF context and using DHS data are scarce, leaving policymakers without localized evidence to guide interventions (13). A part from dietary factors socioeconomic and maternal factors further shape child nutritional outcomes. Maternal education improves awareness of proper feeding practices and health-seeking behaviors, while household wealth affects food affordability and dietary diversity(14). In Tajikistan, rural households sometimes face additional challenges, including poorer maternal education, restricted healthcare access, and inadequate market connectivity (15). These discrepancies contribute to dietary deficiency and greater anemia prevalence among children in impoverished groups. Understanding how socioeconomic disparities connect with ZVF consumption is therefore crucial for devising effective, equitable nutrition programs.

Using data from the 2023 Tajikistan Demographic and Health Survey (TjDHS), this study examines whether complete absence of fruit and vegetable consumption independently associates with anemia among children aged 0-59 months, filling a critical evidence gap in Central Asia. It investigates how this relationship varies by household wealth, maternal factors, and urban-rural residence. Notably, null findings would be equally valuable indicating that dietary interventions alone may be insufficient without addressing upstream determinants such as maternal health status, intrauterine growth, and poverty. The findings are intended to offer evidence based insights that will direct national nutrition plans and inform specific policies there by targeting SDG 2.2 (ending all forms of malnutrition by 2030) and SDG 3.2 (reducing preventable child deaths).

### Conceptual Framework

The conceptual framework for childhood anemia recommended by UNICEF, adapts its broader framework for maternal and child undernutrition. It identifies a hierarchy of causes, including immediate causes like inadequate diet,illness and health care, underlying causes such as household food insecurity and health policies, and basic causes stemming from social, economic, and political factors. UNICEF’s approach emphasizes that interventions must be integrated and addressed at all levels of these determinants(10). Nutritional iron intake is the most immediate or downstream pathway that is responsible for child’s anemia.

## Methodology

### Study Design and Setting

We conducted a cross-sectional study that utilized secondary data analysis from the DHS 2023 Tajikistan, (TjDHS). Thereby we were interested to investigate the association between ZVF consumption with anemia in children aged 0-59 months in this country. The TjDHS,2023 the third following surveys in 2012 and 2017, took place from August to November 2023 during post-harvest months, potentially underrepresenting food shortages common in off-seasons. Tajikistan’s mountainous landscape, reliance on staple crops like rice and wheat, and climate challenges likely heighten child malnutrition and anemia, an ongoing public health issue.

Our study adhered to strengthening the Reporting of Observational Studies in Epidemiology (STROBE) guidelines which emphasized structured overview of design, variables, and analyses enhancing replicability and robust estimates thus minimizing reporting biases (16).

### Sampling Methods and Sample Frame

The TjDHS utilized a two-stage stratified cluster sampling design(17), covering all regions and balancing urban and rural areas to provide a nationally representative profile of child anemia. It involved nine strata (urban/rural across regions). In the first stage, 370 primary sampling units (PSUs) were selected with probability proportional to size, based on the 2020 Tajikistan Population and Housing Census [TPHC] as its sampling frame conducted by the Agency on Statistics under the President of the Republic of Tajikistan (Tajstat). In the second stage, 22 households per PSU were systematically selected from updated household listings, yielding 8,140 households (3,652 urban, 4,488 rural). This design, with no replacements allowed, ensured a representative sample of 10,718 completed interviews with women age 15–49 covering all regions(17,18).(Figure-2)

**FIGURE 1:**
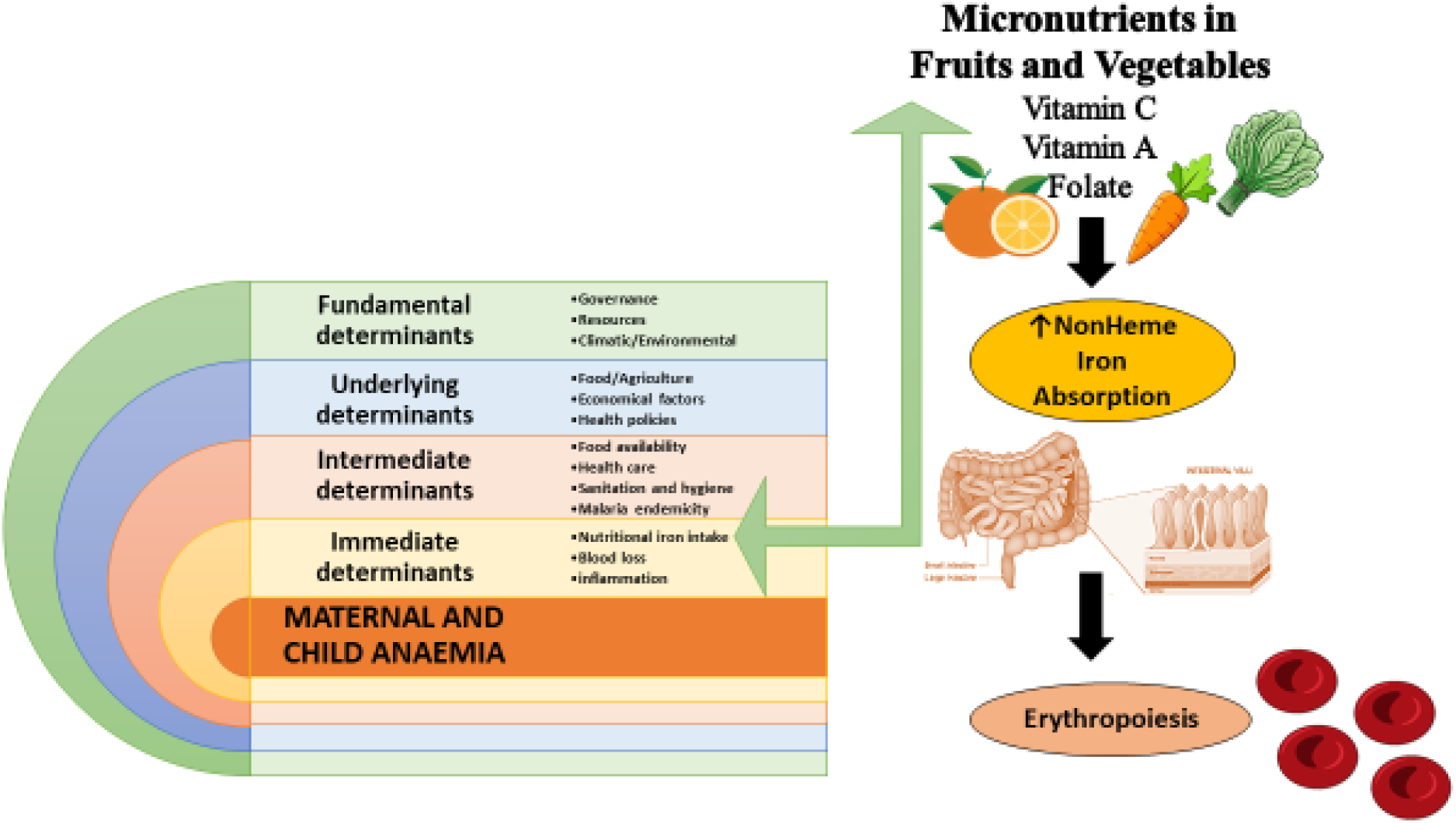
Conceptual Framework of Maternal Child Anemia. zvf folder\CONCEPTUAL FRAMEWORK ZVF-fig 1.pptx Source: Unicef^8^

**Figure 2.**
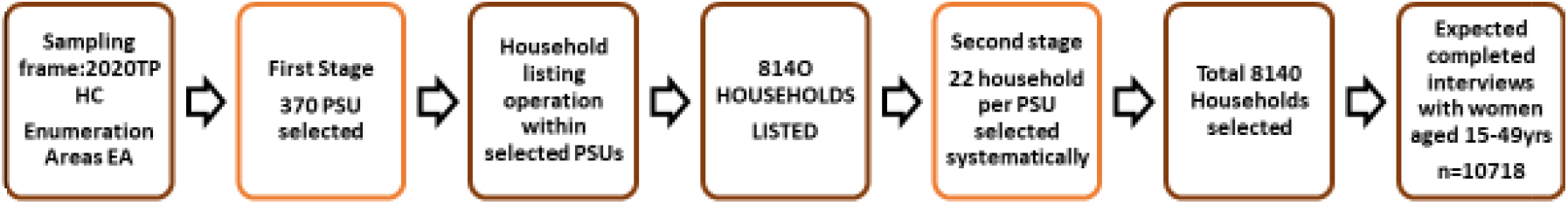
Sampling Frame. zvf folder\Sampling Frame-fig 2.pptx Source: TjDHS-2023

Where,

TPHC- Tajikistan Population and Housing Census

PSU- Primary Sampling Units

### Study Participants and Eligibility

The target population comprised children with age 0–59 completed months and non-pregnant women 15-49 completed years of age, with no distinction between lactating and non-lactating women residing in households selected for the 2023 DHS survey in Tajikistan. This total reflects all de jure participants. All eligible children in these households were recruited, minimizing selection bias(18). Inclusion criteria included children with complete data on hemoglobin levels and dietary intake in the preceding 24 hours. Children over 59 months (per WHO anemia guidelines) were excluded. The analytic sample consisted of 2,355 children after exclusions for missing data on specific variables (e.g. child age, anemia, dietary variables) representative of the 2023 Tajikistan DHS, which surveyed 8,035 households with a 99.6% response rate, covering all regions and urban-rural strata. The survey weights minimized non-response bias and ensured representativeness(18).

### Data Collection

Data was collected via standardized DHS questionnaires involving the Woman’s Questionnaire that collected information from all eligible women age 15-49yrs related to health and nutrition, breastfeeding and complementary feeding practices. The Biomarker Questionnaire was used to record the results of anemia testing. The protocol for anemia testing was reviewed and approved by the Institutional Review Board of Ministry of Health and Social Protection of the Population of Tajikistan (MoHSPP) and the ICF, UNICEF and Asian development bank(17). The DHS surveys use single method, the HemoCue system for hemoglobin measurement.

### Quality Assurance

Hemoglobin measured by trained enumerators following DHS protocols, with quality control Blood specimens for anemia testing were collected from women age 15-49yrs who consented to be tested and from children age 0–59 months whose parents or guardians had given consent to the testing. Capillary blood was drawn from a finger prick (or a heel prick in the case of children age 6–11 months and the third drop of capillary blood was collected in a HemoCue® 201+ microcuvette. Hemoglobin analysis was carried out on-site using a battery-operated portable HemoCue® 201+ device, daily HemoCue device calibration was also checked. Altitude adjustment applied using DHS-provided geospatial data. Hemoglobin levels were measured following established Demographic and Health Surveys (DHS) protocols, utilizing trained enumerators to ensure data consistency and accuracy. To maintain data reliability and validity, a rigorous quality control process was implemented, including duplicate testing on a 5% subsample of measurements. Furthermore, all hemoglobin values were adjusted for altitude using geospatial data provided by DHS, a critical step to account for physiological variations at different elevations. Field supervision by team of supervisors, field coordinators and biomarker specialists observed procedures directly and use checklists ensure accurate measurements, achieving ∼95% biomarker coverage(17). Any necessary data cleaning steps were performed to enhance the integrity of the dataset.

### Completeness of Data

A precise analysis of 2023 data survey of Tajikistan was done by a team of researcher A.K and B.A, in order to validate the completeness of data for children aged 0–59 months. The data showed a total of 5,068 cases with 135 missing data for child’s age. A stepwise approach was done to handle the missing values by listwise deletion from the system. Next the data on child’s anemia was checked for appropriateness. This included 413 “Missing” cases as explicitly and “System” missing (likely due to incomplete or unrecorded hemoglobin measurements). Among the valid cases, 4520 children had complete anemia data. No specific outliers were reported during data cleaning suggesting values were within expected ranges after quality control by trained field staff and biomarker monitors (∼95% biomarker coverage). The 413 missing cases were excluded by listwise deletion from data finalizing complete anemia data. Further missing values were similarly handled from various food diversity variables thus ensuring completeness of data of valid 2355 cases.(Figure-3)

**Figure 3:**
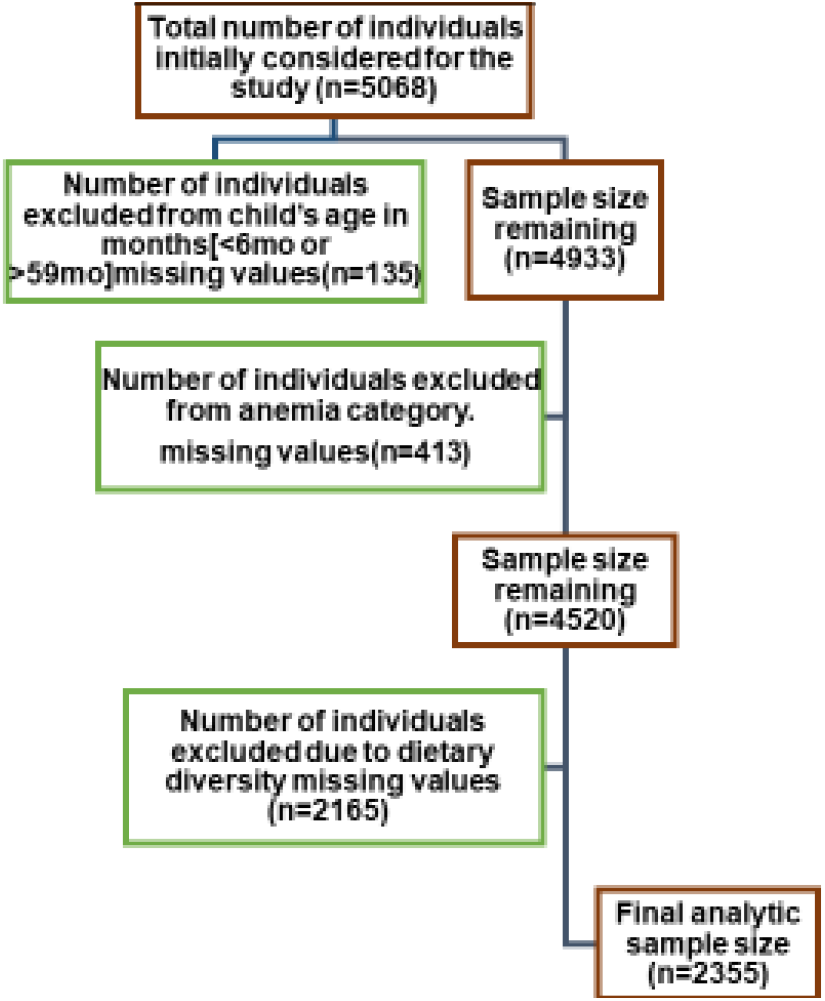
Strobe flow diagram for participants and eligibility criteria for DHS analysis. zvf folder\Strobe flow diagram-fig 3.pptx

### Study Variables

#### Exposure Variables

##### Zero Vegetable or Fruit consumption:[ZVF]

ZVF consumption 0-59 months reflects percentage of children age 0-59 months who were not fed any vegetables or fruits during the previous day. Number of youngest children age 0-59 months living with their mother(8). In the 2023 Tajikistan Demographic and Health Survey (TjDHS), data on vegetable and fruit consumption for children aged 0-59 months were gathered via structured 24-hour dietary recall interviews with mothers or primary caregivers, using a standardized questionnaire. Fieldworkers queried whether children consumed specific items in the past day, including fruit juice (V410), other vegetables (V414A), plantains/potatoes/cassava (V414F), pumpkin/carrots/squash (V414I), dark green leafy vegetables (V414J), mangoes/papayas/vitamin A-rich fruits (V414K), and other fruits (V414L), ensuring consistent and reliable dietary assessment. Responses were recorded as binary (yes/no) for each food item, later combined into a derived variable, ZVF Consumption (ZVF), Children who responded “no” to all seven vegetable and fruit consumption variables were classified as having zero consumption (ZVF=1), while those who consumed at least one item were classified as having consumed vegetables and fruits (ZVF=0), (19) [Supplementary file 1].

##### Minimal meal frequency: [MMF]

Minimum meal frequency is a proxy for meeting energy requirements. Breastfed children age 6–8 months are considered to be fed with a minimum meal frequency if they receive solid, semisolid, or soft foods at least twice a day. Breastfed children age 9–23 months are considered to be fed with a minimum meal frequency if they receive solid, semisolid, or soft foods at least three times a day. Non breastfed children age 6–23 months are considered to be fed with a minimum meal frequency if they receive solid, semisolid, or soft foods or milk feeds at least four times a day and if at least one of the feeds is a solid, semisolid, or soft food(8).

### Outcome Variable

#### Child Anemia

The children presented in this study used the new cutoffs to define anemia and have been adjusted for altitude according to the latest WHO guidance. Anemia is defined as hemoglobin <10.5g/dl for 6-23mo and <11 g/dl for 24-59mo, adjusted for altitude per WHO standards to account for reduced oxygen availability in Tajikistan’s mountainous regions. Anemia levels were categorized as severe, moderate, or mild using hemoglobin concentrations based on World Health Organization criteria. Severe anemia was defined as hemoglobin below 7.0 g/dL, moderate anemia as 7.0–9.9 g/dL, and mild anemia as 10.0 g/dL up to the age-specific cutoff (20)(Table 1), ensuring standardized classification for analysis. The categories of anemia severe=1, moderate=2 and mild=3 were merged and coded as “with anemia” new code as 1 and the category not anemic = 4 was coded as 0 “no anemia”. This coding resulted the formation of a binary outcome variable. At the time of survey data collection, single-drop capillary blood was still recommended and was used to measure hemoglobin(21).

**Table 1.**
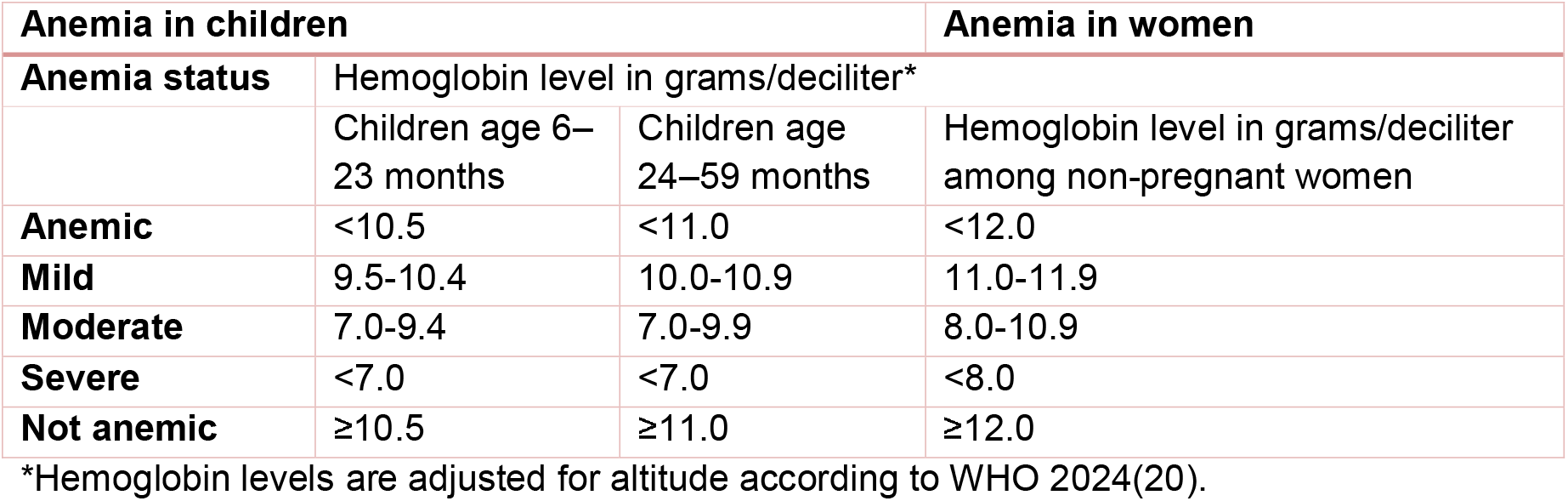
Anemia cutoff points in accordance with WHO 2024. zvf folder\Table 1-ZVF.docx

### Covariates

Covariates were chosen based on prior Demographic and Health Survey research on child anemia, targeting socioeconomic, demographic, and health-related factors linked to nutritional deficiencies in low- and middle-income settings. These included child age (6–23 months for early feeding risks, 24–59 months for high body demands), sex to address potential gender-based feeding variations, urban-rural residence to account for food and healthcare access gaps, maternal education (none, primary, secondary+) as a marker of health knowledge, household wealth index from asset-based analysis to reflect economic limits, birth order for household resource distribution, recent morbidity (diarrhea or fever in past two weeks) affecting nutrient uptake, and minimum meal frequency to control for feeding frequency’s impact on nutrient intake in children aged 6–59 months.

### Ethical Considerations

This secondary analysis used de-identified public data hence no additional ethical approval required. Original DHS adhered to Helsinki Declaration ethical framework for the original survey’s data collection, ensuring informed consent was obtained from the participants of the study.

### Statistical Analysis

Descriptive statistics characterized the study population using weighted frequencies and percentages for categorical variables were utilized. Inferential analyses utilized binomial and multivariable logistic regression, conducted in Jamovi version 2.3 and SPSS version 25 softwares, to estimate unadjusted and adjusted odds ratios (ORs) with 95% confidence intervals for the association between ZVF intake and anemia among children aged 0-59 months. We used binomial logistic regression to examine factors associated with childhood anemia, treating anemia status as a binary outcome (anaemic vs. non-anaemic). The analysis proceeded in two stages. First, we ran separate unadjusted models for each predictor variable to identify their crude associations with anemia. Variables that showed statistical significance (p < 0.05) in these univariate analyses were then carried forward into a multivariable model.The final multivariable logistic regression model included ZVF consumption (ZVF1), meal frequency (M39), household wealth index (V190), maternal anemia status (V457A), and perceived birth size (M18). We reported odds ratios (OR) with 95% confidence intervals to quantify the strength and direction of associations. Model fit was evaluated using McFadden’s R^2^, chi-square tests, and information criteria (AIC and BIC). To check for multicollinearity among predictors, we calculated variance inflation factors (VIF) and tolerance statistics, with VIF values below 5 considered acceptable.All analyses accounted for the complex survey design and sampling weights of the DHS data to ensure nationally representative estimates. Statistical analyses were conducted using jamovi (Version 2.3) with Statistical Package for Social Sciences(SPSS Version 25). Statistical significance was set at p < 0.05 for all tests.

## Results

### Sociodemographic and Health Characteristics of the Study Population

The study included 2,355 children aged 0-59 months, with a nearly equal distribution between males (50.4%) and females (49.6%). The largest proportion of children were in the 12-23 months age group (39.2%), followed by infants aged 0-11 months (21.7%), while children aged 24-35 months, 36-47 months, and 48-59 months constituted 12.2%, 14.6%, and 12.4% of the sample, respectively. The vast majority of children were singleton births (98.3%), with only 1.7% being twins or triplets. Among those with recorded birth size data (n=1,720), approximately two-thirds (65.7%) were perceived as average size at birth, 23.5% as large, and 10.8% as small. Birth weight information was available for 1,720 children, with 95.1% having normal birth weight and 4.9% classified as low birth weight. Regarding birth order, 63.9% were subsequent children while 36.1% were index children. Recent childhood morbidities showed that 18.0% of children experienced diarrhea in the two weeks preceding the survey, 11.0% had fever, and 4.2% presented with symptoms of acute respiratory infection characterized by short, rapid breaths. Maternal characteristics revealed that over half of the mothers (54.6%) were aged 25-34 years, followed by those aged 15-24 years (35.6%) and 35 years or older (9.7%). Maternal nutritional status showed considerable variation, with 38.0% having normal BMI, 36.0% being overweight, 20.6% obese, and 5.5% underweight. Maternal anemia was also highly prevalent, affecting 34.6% (n=815) of mothers, while 65.4% (n=1,538) were not anaemic. Educational attainment indicated that three-quarters of mothers (75.1%) had lower education levels, while 24.9% had higher education. At the household level, the majority of families were of medium size with 6-10 members (59.5%), followed by small households with 0-5 members (21.1%) and large households with more than 10 members (19.4%). The sample was predominantly rural (60.8%) compared to urban (39.2%). Wealth distribution showed relative heterogeneity, with the richest index representing the largest group (29.9%), while the poorest, poorer, middle, and richer indexs accounted for 16.9%, 17.2%, 17.7%, and 18.4% of the sample, respectively.(Table-2)

### Prevalence of Childhood Anemia, Feeding Frequency and ZVF Consumption

The overall prevalence of anemia among children under five years in Tajikistan was 33.2% (n=782), indicating that approximately one-third of the study population was anaemic, while 66.8% (n=1,573) were non-anaemic. Analysis of child feeding practices revealed considerable variation in meal frequency among children. Among those for whom feeding data were available (n=1,329), the majority of children consumed solid, semi-solid, or soft foods twice daily (47.3%), followed by once daily (23.7%), while 16.4% reportedly consumed no such foods in the previous 24 hours. Smaller proportions consumed these foods three times (9.1%), four times (1.8%), five times (1.2%), sxsix times (0.4%), or seven times (0.2%) daily, with 1.4% of caregivers unable to recall feeding frequency. Critically, the derived variable for ZVF consumption indicated that 36.1% (n=851) of children consumed no vegetables or fruits whatsoever in the 24 hours preceding the survey, while 63.9% (n=1,504) consumed at least one type of vegetable or fruit.(Figure-4)

Where, ZVF=Zero Vegetable or Fruit consumption.

**Figure 4.**
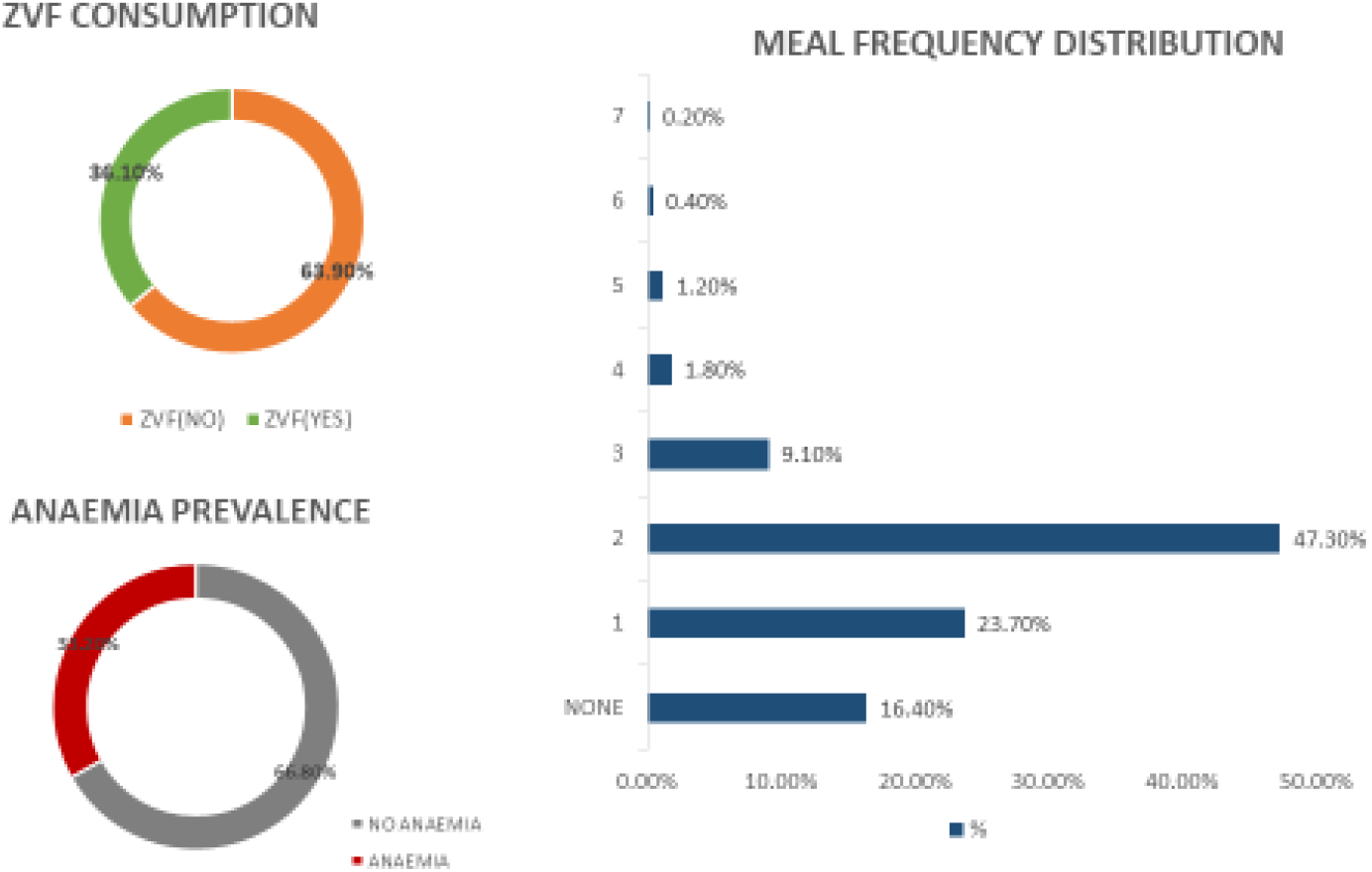
Prevalence OF ZVF, Child anemia, Minimum Meal Frequency. zvf folder\Prevalence of ZVF,anaemia,MMF-fig 4.pptx

### Association Between ZVF Consumption and Childhood Anemia

In the unadjusted binomial logistic regression analysis from our study, ZVF consumption showed no statistically significant association with childhood anemia. However only marginally higher but non-significant odds of anemia were revealed when compared to those who consumed vegetables and fruits (OR = 1.03, 95% CI: 0.86 to 1.24), indicating no crude association between dietary diversity and anemia status. However, when ZVF consumption was examined in the multivariable adjusted model that controlled for months since last birth (M39), wealth index (V190), maternal anemia status (V457A), and perceived birth size (M18), the direction of association reversed, though it remained statistically non-significant that is had lower odds of anemia compared to those who consumed vegetables and fruits (OR = 0.85, 95% CI: 0.63 to 1.14), representing an approximate 15% non-significant reduction in anemia odds. The multivariable model demonstrated good overall fit [(chi square)χ^2^ = 69.7, (degree of freedom)df = 9, p < 0.001, (model fit) McFadden’s R^2^ = 0.0398) and satisfied collinearity assumptions with all variance inflation factors remaining below 1.20, confirming the stability of parameter estimates.

### Association of Significant Covariates with Childhood Anemia

Several maternal, socioeconomic, and birth-related factors demonstrated significant associations with childhood anemia in unadjusted analyses, and these relationships persisted with varying magnitudes after adjustment in the multivariable model. Household wealth index exhibited a strong dose-response relationship with childhood anemia in both unadjusted and adjusted analyses. In the crude model, wealth status was highly significant [χ^2^ = 34.6, df = 4, p < 0.001], with children from the richest index showing approximately 50% lower odds of anemia compared to the poorest index (OR = 0.50, 95% CI: 0.38 to 0.65). After adjusting for other covariates, this protective effect was amplified, with the richest index demonstrating a 60.5% reduction in anemia odds (OR = 0.39, 95% CI: 0.27 to 0.56). Similarly, children from richer households showed a significant, protective effect in crude analysis (OR = 0.73, 95% CI: 0.55 to 0.97), which became stronger in the adjusted model (OR = 0.53, 95% CI: 0.36 to 0.78), indicating a 46.2% reduction in anemia odds. The poorer and middle indexes, which showed non-significant protective trends in unadjusted models, maintained similar non-significant associations after adjustment.

Maternal anemia status emerged as a critical predictor of childhood anemia in both analytical approaches. In unadjusted analysis, maternal anemia was significantly associated with increased childhood anemia (χ^2^ = 14.4, df = 1, p < 0.001), with children of anemic mothers having 41.4% higher odds of being anaemic (OR = 1.41, 95% CI: 1.18 to 1.68). This intergenerational transmission effect became more pronounced in the adjusted model, where children of anaemic mothers demonstrated 48.1% higher odds of anemia (OR = 1.48, 95% CI: 1.16 to 1.88), suggesting that maternal nutritional status exerts an independent effect on child anemia that is not mediated by socioeconomic conditions or birth characteristics.

Perceived birth size at delivery showed a significant association with anemia in unadjusted analysis (χ^2^ = 11.4, df = 2, p = 0.003), with children perceived as small at birth having substantially elevated odds of anemia compared to average-sized children (OR = 1.68, 95% CI: 1.23 to 2.31). This effect was markedly amplified in the adjusted model, where small birth size was associated with a 92.2% increase in anemia odds (OR = 1.92, 95% CI: 1.31 to 2.81), making it one of the strongest independent predictors of childhood anemia. Large birth size showed no significant association in either unadjusted (OR = 1.18, 95% CI: 0.94 to 1.50) or adjusted (OR = 1.10, 95% CI: 0.83 to 1.44) models, confirming that only growth restriction, not macrosomia, contributes to anemia risk.

Child meal frequency demonstrated a modest but significant protective effect in both models. In crude analysis, each additional meal occasion was associated with an 11.4% reduction in anemia odds (OR = 0.88, 95% CI: 0.79 to 0.98). After controlling for wealth, maternal anemia, birth size, and ZVF consumption, this protective effect remained significant with each additional meal associated with a 13.5% reduction in anemia odds (OR = 0.86, 95% CI: 0.76 to 0.98), underscoring the independent importance of adequate feeding frequency for child health outcomes. The consistency of these effects across unadjusted and adjusted models demonstrates the robustness of meal frequency as a modifiable risk factor for childhood anemia, independent of socioeconomic status and maternal nutritional status.

Several other covariates examined in unadjusted analyses showed no significant associations with childhood anemia. Maternal education level, comparing higher versus lower education (OR = 1.05, 95% CI: 0.86 to 1.28), household size, whether medium (6-10 members) or large (10+ members) compared to small households (0-5 members) (p = 0.191), birth type, comparing twins/triplets to singleton births (OR = 1.16, 95% CI: 0.61 to 2.21) showed no significant association in our study. Child sex was also not significantly associated with anemia, with female children showing marginally lower but non-significant odds compared to males (OR = 0.89, 95% CI: 0.75 to 1.06). Similarly, birth order, comparing subsequent children to index children and reported birth weight, comparing normal to low birth weight demonstrated no significant association in our study. Recent morbidities including fever in the past two weeks, recent diarrhea, and acute respiratory infections all showed no significant associations with childhood anemia status.(Table-3)

## Discussion

Our study’s null finding on the association between ZVF consumption and childhood anemia is novel for Tajikistan and Central Asia, where dietary-anemia links are understudied, with no prior DHS-based analyses focusing on ZVF(13). This result implies that while low vegetable/fruit intake is prevalent (36.1%), anemia in Tajik children is driven more by maternal and socioeconomic factors than this specific dietary element, directing resources toward targeted, non-dietary interventions. There is limited and fragmented studies on the specific association between fruit and vegetable (F&V) intake and anemia in infants and young children, particularly in contexts like rural Tajikistan with ZVF focus. Most available research is cross-sectional from LMICs in Africa (e.g., Uganda, Ethiopia, South Africa) or Asia (e.g., India), with only a handful directly isolating fruits and vegetables (F&V) rather than broader dietary diversity, and even fewer targeting under five years (0-59 months) or rural settings(22). Non-heme iron in F&V offers low direct bioavailability (2-20% absorption) for infants and young children, contributing minimally to iron needs but crucially enhancing absorption from other plant sources via vitamin C, which can boost uptake by 2-6 times countering inhibitors,phytates in cereal-based diets.

Across the studies we reviewed, low F&V consumption is generally associated with higher anemia risk in children, often through reduced vitamin C enhancement of non-heme iron absorption. Studies from Brazil (23) and Uganda (24) link low F&V intake to higher anemia prevalence in children, contrasting our findings of no significant ZVF-anemia association. However, results are mixed, some Ethiopian studies(25) show low or no direct correlation, aligning with our result, suggesting confounding by socioeconomic factors, infections, or staple-heavy diets that overshadow F&V’s indirect benefits. In infant/young child contexts, F&V aren’t strong direct iron sources but help mitigate Iron deficiency anemia IDA via diversity; protective effects are clearer with vitamin C-rich options,supported by an Indian evidence(26) that emphasized vitamin C-rich F&V’s role in enhancing iron absorption, suggesting our Tajik children’s traditional staple-heavy diets may dilute ZVF effects unless combined with bioavailable iron sources. South African data(27) ties low F&V nutrient density to anemia with rural-targeted diversity interventions. The null associations as the study’s case highlight the need for broader interventions beyond ZVF alone, such as addressing maternal anemia or rural access barriers. The protective effect of increased food frequency as revealed from our study is consistent with WHOs recommendations (28) on infant and young child feeding practices suggests that increasing the number of feeding opportunities can have a positive impact on nutritional status even in low-diversity diets potentially compensating for low ZVF consumption(28)(Table 3).

The lack of ZVF-anemia association in our study may stem from methodological, biological, and cultural factors. Methodologically, we combined the three categories of anemia (mild, moderate, and severe) into a binary outcome (anemia vs. no anemia) for this analysis. One major drawback of this aggregation is that it might have obscured the actual effects of ZVF thus resulting in combining the more common, frequently multifactorial mild cases with the less common, nutrient-sensitive severe cases, as suggested by Shibeshi et al. 2024,(26). Biologically, anemia’s high prevalence (33.2%) likely reflects iron deficiency from infections (18% diarrhoea, 11% fever) or non-heme iron diets(63.9%)(29). Culturally, rural Tajik diets rely on high consumption of dairy products and starchy staples and a low intake of meat, eggs, and nutrient-dense foods(30). Thus, inadequate intake of bioavailable iron (from animal sources) and persistent subclinical infections that prevent iron absorption and utilization may be the main causes of the anemia seen(29) thereby suggesting ZVF alone may not address the dominant iron deficiency drivers in this high-burden setting unless it is combined with greater availability and intake of other bioavailable iron sources.

Despite the inconclusive nature of the primary hypothesis in our study, the adjusted model produced a number of extremely significant and robust risk factors for child anemia that are highly consistent with both domestic and international epidemiological research. The most compelling results emphasized the importance of maternal health. Maternal health strongly predicts child anemia in Tajikistan, particularly in rural settings. Children of anaemic mothers (34.6% prevalence; Table 1) faced significantly higher odds of anemia, reflecting intergenerational iron deficiency transmission(31,32). This link is conciliated by inadequate iron stores passed from mother to fetus(33). Specifically, the study by Siddiqui et al. (2024) suggested that maternal anemia during pregnancy adversely affects placental function and leads to decreased neonatal iron stores, which are vital for an infant’s first 4-6 months of life and directly influence early anemia status(33). Underweight mothers also increased child anemia risk, indicating chronic maternal nutritional deficiencies (34).

Younger maternal age (15-24 years) was a risk factor compared to 25-34 years, possibly due to less experience in diverse feeding practices. Low maternal education (75.1% with lower education; Table 1) may further limit awareness of vegetable/fruit benefits in rural areas, where media exposure and extension services are scarce, though this was not significant in adjusted models(35).These findings underscore the need for pre- and peri-conceptional maternal nutrition interventions to improve child outcomes in rural Tajikistan(36).

The null ZVF-anemia association of Tajikistan’s dietary context as interpreted above is linked with socioeconomic context, particularly in rural areas where majority of the children resides(60.8%) and face a higher anemia risk due to limited access to diverse foods and markets, especially during winter. Some studies also highlight that rural areas are also disproportionately affected by low maternal education and economic status(37). This discrepancy is essentially related to access and infrastructure issues. In addition, poor clusters in urban areas also face similar issues with limited access to healthcare, clean water, and sanitation, stressing the point that the problem is systemic and linked to socioeconomic disparities all across Tajikistan(38).

Socioeconomic gradients further amplify this as children in the richest wealth index had significantly two thirds lower anemia odds compared to the poorest, suggesting ZVF may reflect poverty driven dietary constraints rather than directly causing anemia. Analysis of DHS data from several Sub-Saharan African countries, discovered that the household wealth index was repeatedly linked with lower odds of childhood anemia, independent of maternal and child-level factors(39). Large family sizes (19.4% of households with 10+ members; Table 2) in rural areas may further strain food resources, diluting access to perishables like fruits and vegetables, though this showed no significant adjusted association (Table 2). Younger children (12-23 months) and those with small birth size are particularly vulnerable during weaning, when ZVF introduction is critical but often inadequate in rural households(40).

**Table 2.** Demograhic Characteristics of health and Feeding profile of Tajikistan’s infants and young children 0-59mo. zvf folder\Table 2-ZVF.docx

| Variables | Categories | Weighted frequency (N) | Percentage (%) |
| --- | --- | --- | --- |
| **Child Factors** |  |  |  |
| **Child's Age** | 0-11 | 510 | 21.7% |
|  | 12-23 | 922 | 39.2% |
|  | 24-35 | 288 | 12.2% |
|  | 36-47 | 344 | 14.6% |
|  | 48-59 | 291 | 12.4% |
| **Sex** | Male | 1188 | 50.4% |
|  | Female | 1167 | 49.6% |
| **Birth Type** | Singlet | 2314 | 98.3% |
|  | Twin / triplets | 41 | 1.7% |
| **Birth Size** | Large | 405 | 23.5% |
|  | Small | 185 | 10.8% |
|  | Average | 1130 | 65.7% |
| **Birth Weight** | Low birth weight | 84 | 4.9% |
|  | Normal Birthweight | 1636 | 95.1% |
| **Birth Order** | Index Child | 850 | 36.1% |
|  | Subsequent Child | 1505 | 63.9% |
| **Food frequency** | None | 217 | 16.4% |
|  | 1 | 315 | 23.7% |
|  | 2 | 628 | 47.3% |
|  | 3 | 121 | 9.1% |
|  | 4 | 24 | 1.8% |
|  | 5 | 16 | 1.2% |
|  | 6 | 5 | 0.4% |
|  | 7 | 3 | 0.2% |
| **ZVF consumption** | No | 1504 | 63.9% |
|  | Yes | 851 | 36.1% |
| **Shortness of breaths** | No | 2255 | 95.8% |
|  | yes | 98 | 4.2% |
| **Fever** | No | 2094 | 88.9% |
|  | yes | 258 | 11.0% |
| **Diarrhea** | No | 1928 | 81.9% |
|  | Yes, last two weeks | 425 | 18.0% |
| **Child Anemia** | No Anemia | 1573 | 66.8% |
|  | Anemia | 782 | 33.2% |
| **Maternal Factors** |  |  |  |
| **Maternal age** | 15-24 | 839 | 35.6% |
|  | 25-34 | 1287 | 54.6% |
|  | 35- or More | 229 | 9.7% |
| **Maternal BMI** | Normal | 894 | 38.0% |
|  | Underweight | 129 | 5.5% |
|  | Overweight | 848 | 36.0% |
|  | obesity | 484 | 20.6% |
| **Maternal anemia** | Anemia | 815 | 34.6% |
|  | No anemia | 1538 | 65.4% |
| **Maternal education** | Lower education | 1769 | 75.1% |
|  | Higher education | 586 | 24.9% |
| **Household Factors** |  |  |  |
| **Family size** | Small size (0-5) | 496 | 21.1% |
|  | Medium size (6-10) | 1401 | 59.5% |
|  | Large size (10+) | 458 | 19.4% |
| **Type of place of residence** | Urban | 923 | 39.2% |
|  | Rural | 1432 | 60.8% |
| **Wealth index** | Poorest | 398 | 16.9% |
| combined | Poorer | 404 | 17.2% |
|  | Middle | 417 | 17.7% |
|  | Richer | 433 | 18.4% |
|  | Richest | 703 | 29.9% |

**Table 3:**
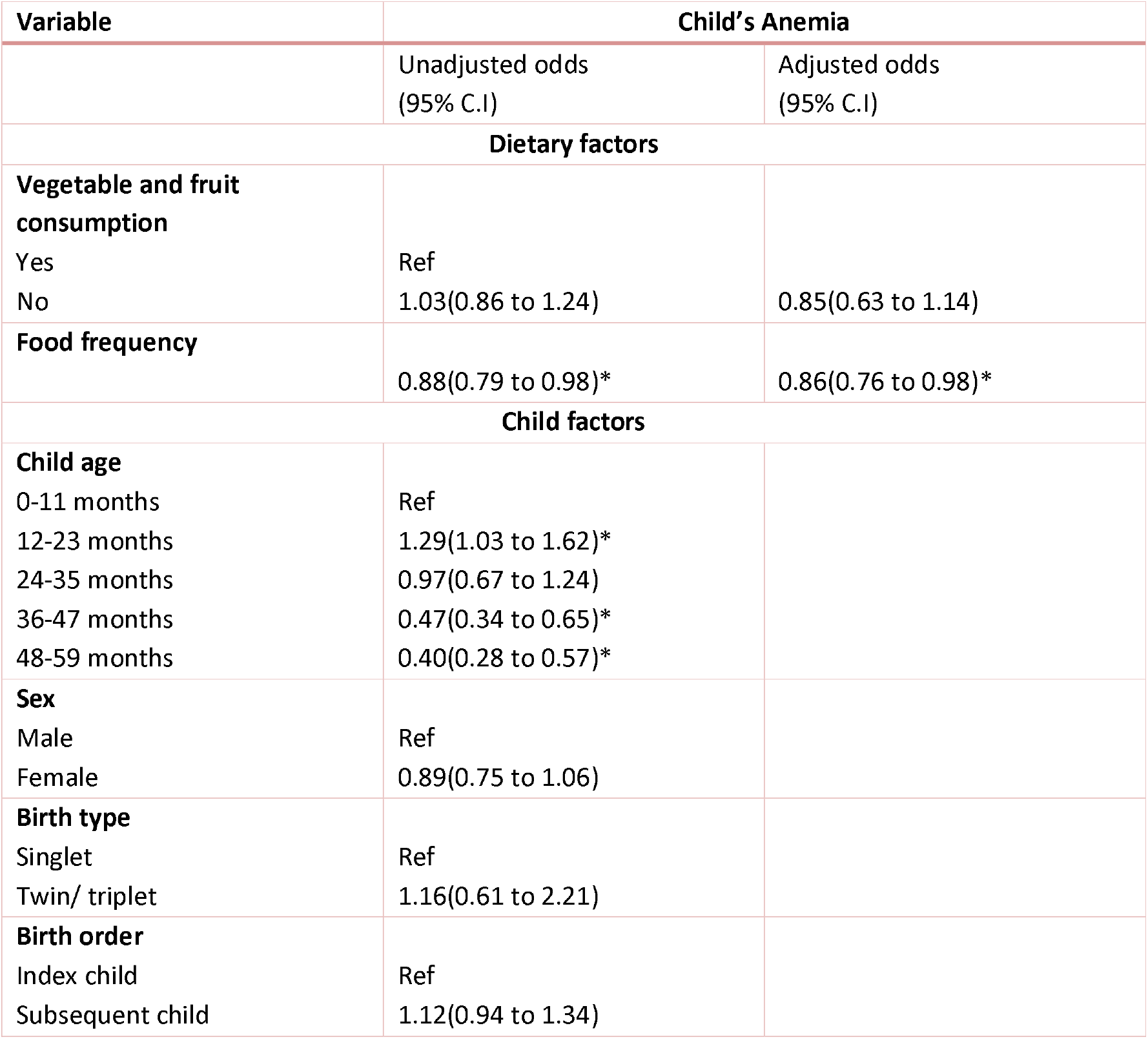
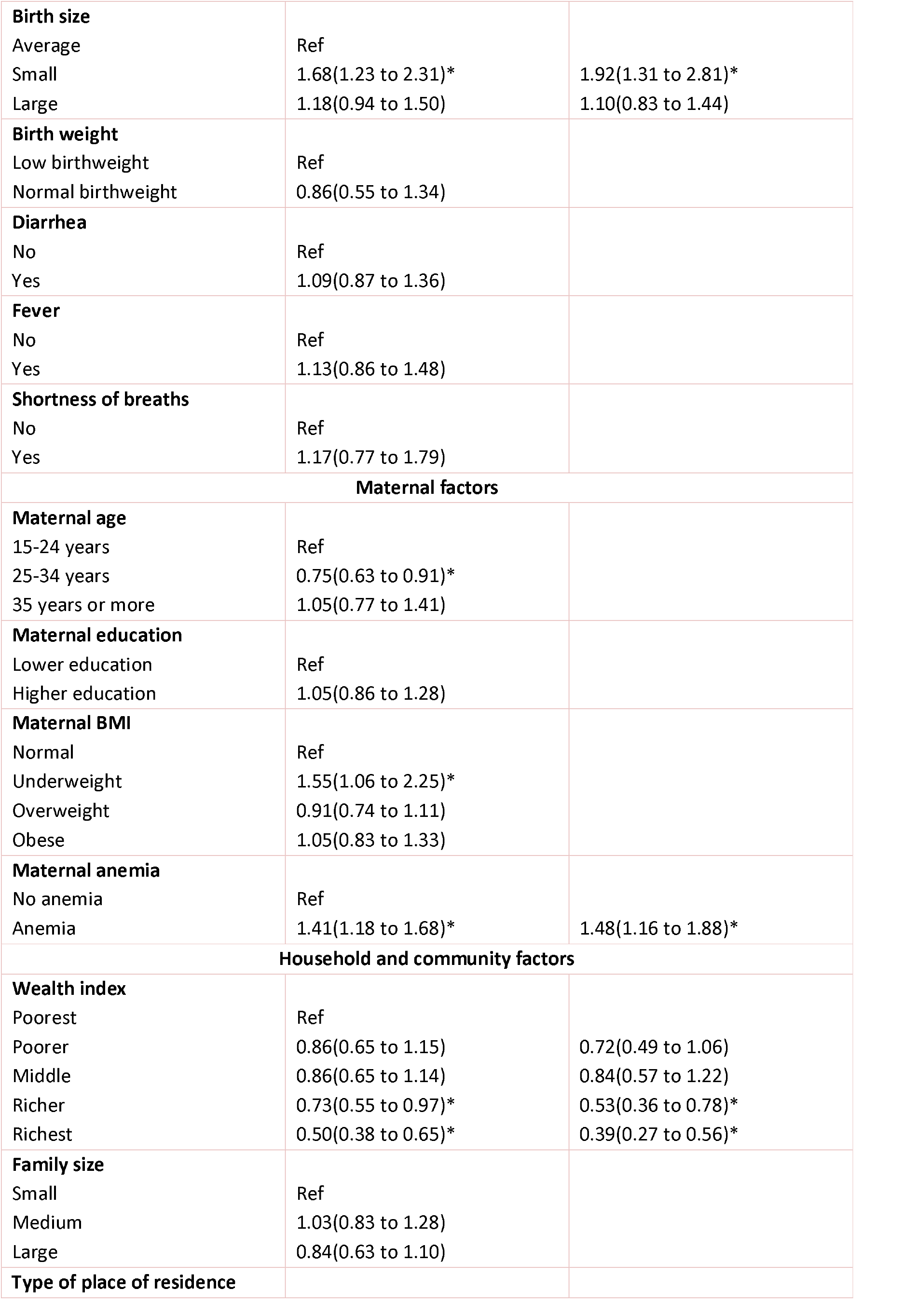
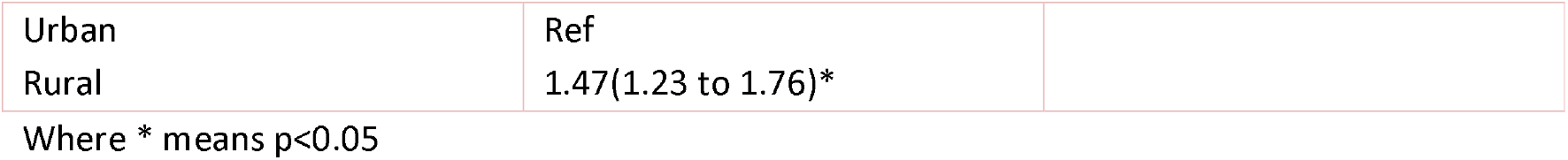
Examining the association of ZVF consumption with anemia among Tajikistan’s infants and young children. zvf folder\Table 3-ZVF.docx

| Variable | Child's Anemia |  |
| --- | --- | --- |
|  | Unadjusted odds<br>(95% C.I) | Adjusted odds<br>(95% C.I) |
| **Dietary factors** |  |  |
| **Vegetable and fruit consumption** |  |  |
| Yes | Ref |  |
| No | 1.03(0.86 to 1.24) | 0.85(0.63 to 1.14) |
| **Food frequency** |  |  |
|  | 0.88(0.79 to 0.98)* | 0.86(0.76 to 0.98)* |
| **Child factors** |  |  |
| **Child age** |  |  |
| 0-11 months | Ref |  |
| 12-23 months | 1.29(1.03 to 1.62)* |  |
| 24-35 months | 0.97(0.67 to 1.24) |  |
| 36-47 months | 0.47(0.34 to 0.65)* |  |
| 48-59 months | 0.40(0.28 to 0.57)* |  |
| **Sex** |  |  |
| Male | Ref |  |
| Female | 0.89(0.75 to 1.06) |  |
| **Birth type** |  |  |
| Singlet | Ref |  |
| Twin/ triplet | 1.16(0.61 to 2.21) |  |
| **Birth order** |  |  |
| Index child | Ref |  |
| Subsequent child | 1.12(0.94 to 1.34) |  |
| **Birth size** |  |  |
| Average | Ref |  |
| Small | 1.68(1.23 to 2.31)* | 1.92(1.31 to 2.81)* |
| Large | 1.18(0.94 to 1.50) | 1.10(0.83 to 1.44) |
| **Birth weight** |  |  |
| Low birthweight | Ref |  |
| Normal birthweight | 0.86(0.55 to 1.34) |  |
| **Diarrhea** |  |  |
| No | Ref |  |
| Yes | 1.09(0.87 to 1.36) |  |
| **Fever** |  |  |
| No | Ref |  |
| Yes | 1.13(0.86 to 1.48) |  |
| **Shortness of breaths** |  |  |
| No | Ref |  |
| Yes | 1.17(0.77 to 1.79) |  |
| **Maternal factors** |  |  |
| **Maternal age** |  |  |
| 15-24 years | Ref |  |
| 25-34 years | 0.75(0.63 to 0.91)* |  |
| 35 years or more | 1.05(0.77 to 1.41) |  |
| **Maternal education** |  |  |
| Lower education | Ref |  |
| Higher education | 1.05(0.86 to 1.28) |  |
| **Maternal BMI** |  |  |
| Normal | Ref |  |
| Underweight | 1.55(1.06 to 2.25)* |  |
| Overweight | 0.91(0.74 to 1.11) |  |
| Obese | 1.05(0.83 to 1.33) |  |
| **Maternal anemia** |  |  |
| No anemia | Ref |  |
| Anemia | 1.41(1.18 to 1.68)* | 1.48(1.16 to 1.88)* |
| **Household and community factors** |  |  |
| **Wealth index** |  |  |
| Poorest | Ref |  |
| Poorer | 0.86(0.65 to 1.15) | 0.72(0.49 to 1.06) |
| Middle | 0.86(0.65 to 1.14) | 0.84(0.57 to 1.22) |
| Richer | 0.73(0.55 to 0.97)* | 0.53(0.36 to 0.78)* |
| Richest | 0.50(0.38 to 0.65)* | 0.39(0.27 to 0.56)* |
| **Family size** |  |  |
| Small | Ref |  |
| Medium | 1.03(0.83 to 1.28) |  |
| Large | 0.84(0.63 to 1.10) |  |
| **Type of place of residence** |  |  |
| Urban | Ref |  |
| Rural | 1.47(1.23 to 1.76)* |  |

Where \* means $p < 0.05$

Our analysis established that the youngest children (12-23months) are the most vulnerable, with anemia odds declining significantly for children aged 36 months and older. The period of rapid growth and the shift from reliance on maternal iron stores to supplemental feeding are the reasons for this age-related vulnerability, which has been extensively documented(40–42). Small birth size was also a significant risk factor, which is in line with research showing that low birth weight and poor fetal growth are associated with lower iron endowment and a higher chance of iron deficiency anemia later in infancy(43,44).

### Strengths and Limitations

The main advantage of this study is that it employs the nationally representative DHS dataset, which enables the findings to be generalized to Tajikistan’s entire infant and young child population. Additionally, multivariable logistic regression concurrently controls for a myriad of potential confounders. The main limitation is that the cross-sectional nature of the data prevents the inference of causality. As crucially discussed before, the aggregation of all anemia severity levels may have masked the true relationship between specific dietary deficiencies and the more severe forms of the condition thus limiting generalizability. Finally, the dietary data relies on maternal recall, which is subject to recall bias.

## Conclusions

This study examined associations between ZVF consumption and anemia among 2,355 children aged 0-59 months in Tajikistan using 2023 Demographic and Health Survey data. The null finding of our study that is no significant ZVF-anemia association despite 36.1% prevalence of children with zero consumption is methodologically robust and regionally novel, representing the first nationally representative examination of this relationship in Central Asia. This challenges dietary centric approaches common in LMIC nutrition programs. Instead, maternal(intergenerational) and socioeconomic factors emerged as primary drivers, maternal anemia increased child anemia odds by 48%, small birth size nearly doubled risk, and wealth disparities showed pronounced gradients with the richest index having 60% lower anemia odds than the poorest. These findings indicate that effective childhood anemia reduction in Tajikistan and potentially across Central Asia requires SDG aligned integrated interventions addressing maternal nutrition during preconception and pregnancy, improved antenatal care to prevent intrauterine growth restriction, poverty alleviation strategies enhancing food security, and promotion of appropriate feeding frequency.

### Future Research

Future research should employ longitudinal designs and comprehensive dietary assessments to elucidate complex interactions between dietary quality, infectious diseases, and socioeconomic determinants in childhood anemia etiology.

### Policy Implications

Our findings provide actionable, evidence-based guidance for national nutrition programs working toward SDG targets in Central Asia and similar LMIC contexts. The Immediate Priority Actions must be given to maternal anemia screening and treatment in all antenatal care units, with free iron-folic acid supplementation for pregnant and lactating women there by Contributing to SDG 3.1(maternal health), 3.2(child health). This must be paralled with targeted cash or food vouchers transfer for the poorest wealth index thereby contributing to SDG 1.2(poverty reduction), 2.1 (food security). Optimal feeding frequency promotion must be done by Community health worker particularly in rural areas which contribute to SDG 2.2(malnutrition).In the intermediate term enhanced antenatal care quality with ultrasound access and fetal growth monitoring to identify and manage intrauterine growth restriction may require equipment investments and sonographer training in rural health facilities. This *c*ontributes to SDG 3.1 (maternal mortality reduction), 3.8 (universal health coverage). In the long term agricultural diversification policies must be employed beyond staple grains thus contributing to SDG 2.3 (agricultural productivity), 2.4 (sustainable food production). As maternal education and wealth showed protective effects, long-term investment can be done for next-generation outcomes.

## Supporting information

zvf folderSupplementary file.docx

## Abbreviations

ZVF: ZVF intake
df: Degree of Freedom
X^2^: Chi square
CI: confidence interval
OR: odds ratio
MMF: minimum meal frequency
MRM: Micronutrient Related Malnutrition
SDGs: Sustainable Development Goals
WHO: World Health Organization
UNICEF: United Nations International Children Emergency Fund
NNS: National Nutrition Survey
PSU: Primary Sampling Units
EB: Enumeration Blocks
LMICs: Lower- and Middle-Income Countries.
CI: Confidence Interval
OR: Odds Ratio
AIC: Akaike Information Criteria
BIC: Bayesian Information Criteria
VIF: Variance Inflation Factors

## Declarations/Statements

### Authors’ contribution

AK: Conceptualization, data management, data analysis, reviewing, editing, and supervision. NI: Introduction writing and helped in reviewing of manuscript. YR: Discussion writing and helped in reviewing of manuscript. SA: Result writing and helped in reviewing and editing. BA: Original draft writing,methodology section, visualization, and formatting of the manuscript also assisted AK in data analysis and in final review of the manuscript.

### Ethics approval

In this study, an ethical approval waiver was granted because the data used was deidentified, and the research team had no direct contact with the study population. Consequently, individual participant consent and ethical approval of this study was not required. However, the data in each DHS was collected following ethical principles of Helsinki’s declaration.

### Consent to participate

In this study, the research team utilized de-identified data obtained from the DHS repository. This data does not contain any information that could be used to identify or trace individual participants. Consequently, individual participant consent was not required by the primary research team for this study.

### Consent for publication

In this research, the DHS data of Tajikistan was used. To access this data, a formal application was submitted, which included our intention to publish the findings. The DHS data archivist provided us with a letter granting permission to publish our research, with the condition that we acknowledge the DHS data repository in the acknowledgments section of the publication

### Data Availability

The data that support the findings of this study are openly available in The DHS Program at https://dhsprogram.com/.

## Acknowledgement

We would like to acknowledge the data archivist of the Demographic and Health Surveys [DHS] Program, who provided access to the dataset of DHS implemented in Tajikistan for analysis.

## Supplementary file

The supplementary file has two tables, which presents the individual variables of fruit and vegetables and derived variable from them. zvf folder\Supplementary file.docx

---

**Supplementary file 1:**
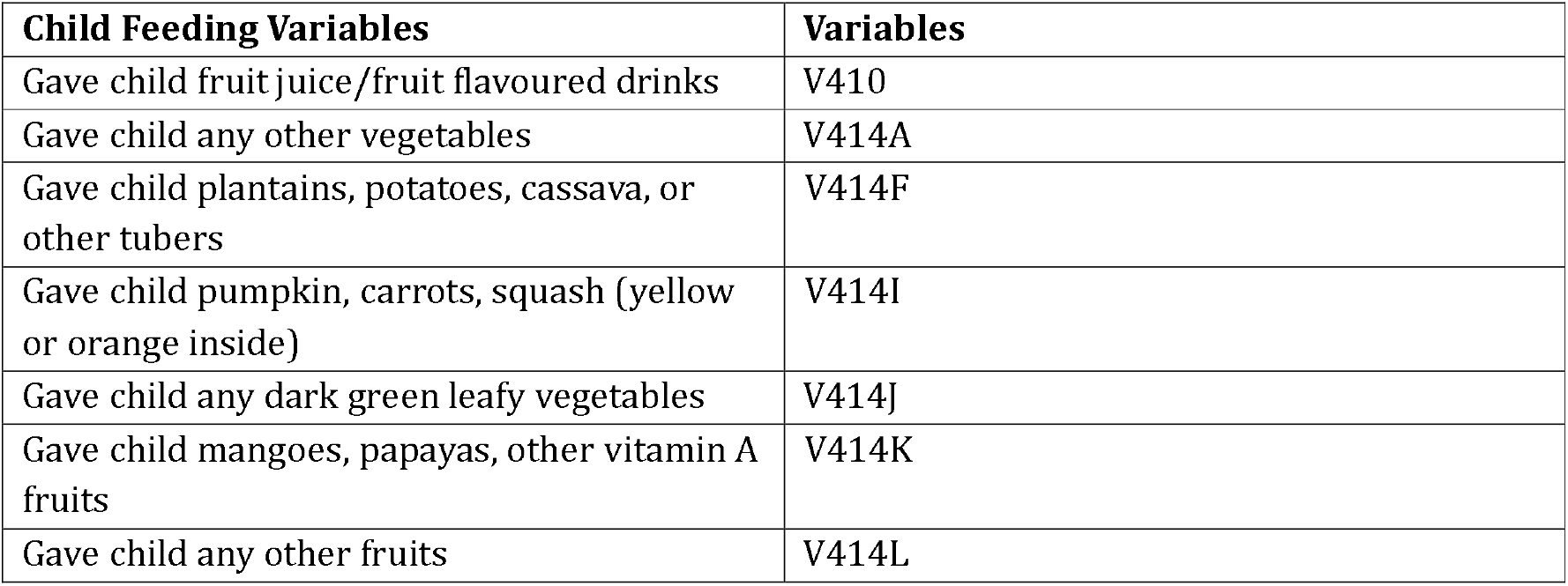
Feeding Variables for infants and child (6-59mo) from DHS Tajikistan 2023.

**Supplementary file 2:**
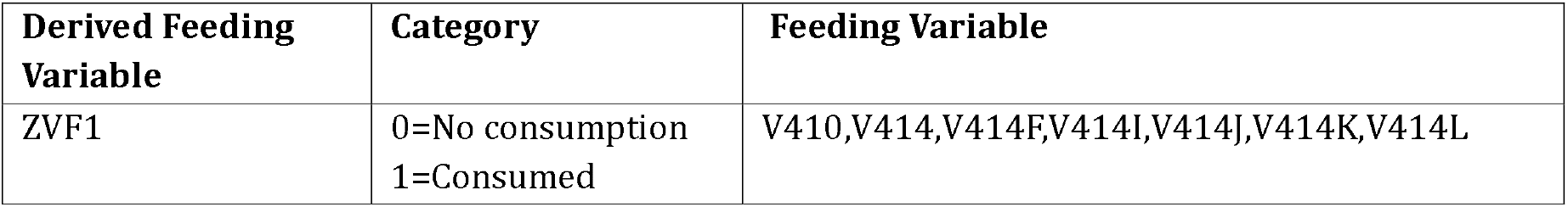
Derived Feeding Variable for infants and child (6-59mo) from DHS Tajikistan 2023.

