## Supplementary material for "“Maternal and Socioeconomic Factors Drive Childhood Anemia in Tajikistan: Examining the Role of Zero Vegetable or Fruit Consumption among under five”": zvf folderSupplementary file.docx

**Supplementary file 1: Feeding Variables for infants and child (6-59mo) from DHS Tajikistan 2023**

| **Child Feeding Variables** | **Variables** |
| --- | --- |
| Gave child fruit juice/fruit flavoured drinks | V410 |
| Gave child any other vegetables | V414A |
| Gave child plantains, potatoes, cassava, or other tubers | V414F |
| Gave child pumpkin, carrots, squash (yellow or orange inside) | V414I |
| Gave child any dark green leafy vegetables | V414J |
| Gave child mangoes, papayas, other vitamin A fruits | V414K |
| Gave child any other fruits | V414L |

**Supplementary file 2: Derived Feeding Variable for infants and child (6-59mo) from DHS Tajikistan 2023**

| **Derived Feeding Variable** | **Category** | **Feeding Variable** |
| --- | --- | --- |
| ZVF1 | 0=No consumption  1=Consumed | V410,V414,V414F,V414I,V414J,V414K,V414L |
